# Age- and Sex-Specific Prevalence of Lower Urinary Tract Symptoms in Pediatric and Young Adults in a Swedish National Cohort

**DOI:** 10.1101/2025.03.15.25324030

**Authors:** Lotta Renström-Koskela, L Viktor Skokic, Adrianna P. Kępińska, Danielle Scharp, Christina Gustavsson Mahjani, Keith Humphreys, Joseph D. Buxbaum, Dorothy E. Grice, Behrang Mahjani, Olof Akre

**Affiliations:** Department of Molecular Medicine and Surgery, Karolinska Institutet, Stockholm, Sweden; Department of Pelvic Cancer, Karolinska University Hospital, Stockholm, Sweden; Seaver Autism Center for Research and Treatment, Icahn School of Medicine at Mount Sinai, New York, NY, USA; Department of Psychiatry, Icahn School of Medicine at Mount Sinai, New York, NY, USA; Department of Genetics and Genomic Sciences, Icahn School of Medicine at Mount Sinai, New York, NY, USA; Division of General Internal Medicine, Icahn School of Medicine at Mount Sinai, New York, NY, USA; Department of Medical Epidemiology and Biostatistics, Karolinska Institutet, Stockholm, Sweden; Mindich Child Health and Development Institute, Icahn School of Medicine at Mount Sinai, New York, NY, USA; Friedman Brain Institute, Icahn School of Medicine at Mount Sinai. New York, NY, USA; Department of Artificial Intelligence and Human Health, Icahn School of Medicine at Mount Sinai, New York, NY, USA

## Abstract

**Background:** Lower Urinary Tract Symptoms (LUTS significantly impact quality of life and are prevalent across life course. Despite their importance, population-based incidence rates for pediatric and young adult populations, including age and sex-specific differences, remain poorly quantified.

**Methods:** We analyzed data from all live-born children in Sweden between 1995 and 2005, with follow-up through 2018, extracted from the Swedish National Medical Birth Register. We employed three distinct methods to identify LUTS cases: (1) an ICD-10 diagnostic code approach, (2) a broad ICD-medication approach that includes ICD-10 diagnosis or at least one prescription of LUTS-related medications (anticholinergics, Alfa-1-receptor antagonist, or Beta-3 receptor agonists), and (3) a narrow ICD-medication approach that includes an ICD-10 diagnosis or multiple prescriptions over a specified timeframe. We analyzed sex-specific (sex assigned at birth) and age-specific incidence rates per 100,000 person-years in individuals younger than 24.

**Findings:** The cohort included 982,903 individuals, 0.1% diagnosed with storage and voiding LUTS based on ICD-10 diagnostic code. Under the broad ICD-medication definition, 0.9% had storage LUTS, and 0.2% had voiding LUTS; the narrow ICD-medication definition identified 0.5% with storage LUTS and 0.1% with voiding LUTS. Notably, females were diagnosed at older ages.

**Conclusions:** This study reveals an underestimation of LUTS prevalence when relying solely on ICD-10 diagnostic codes. Broader diagnostic criteria uncover higher prevalences and demonstrate varied detection rates and ages at diagnosis by sex. These findings underscore the necessity for more inclusive diagnostic approaches in epidemiological research to capture and understand the full scope of LUTS accurately.

## 1. Introduction

Lower Urinary Tract Symptoms (LUTS) are increasingly recognized as a significant public health concern due to their widespread prevalence and substantial impact on an individuals’ quality of life ^1–3^. LUTS manifest a variety of clinical symptoms and signs that are typically divided into three main categories: storage, voiding, and post-micturition (Figure 1). Storage signs include urinary frequency, urgency, nocturia, and various forms of incontinence. Voiding signs are characterized by hesitancy, intermittency, slow stream, and straining during urination. Post-micturition signs generally involve the feeling of incomplete bladder emptying and post-void dribbling ^4^.

**Figure 1.**
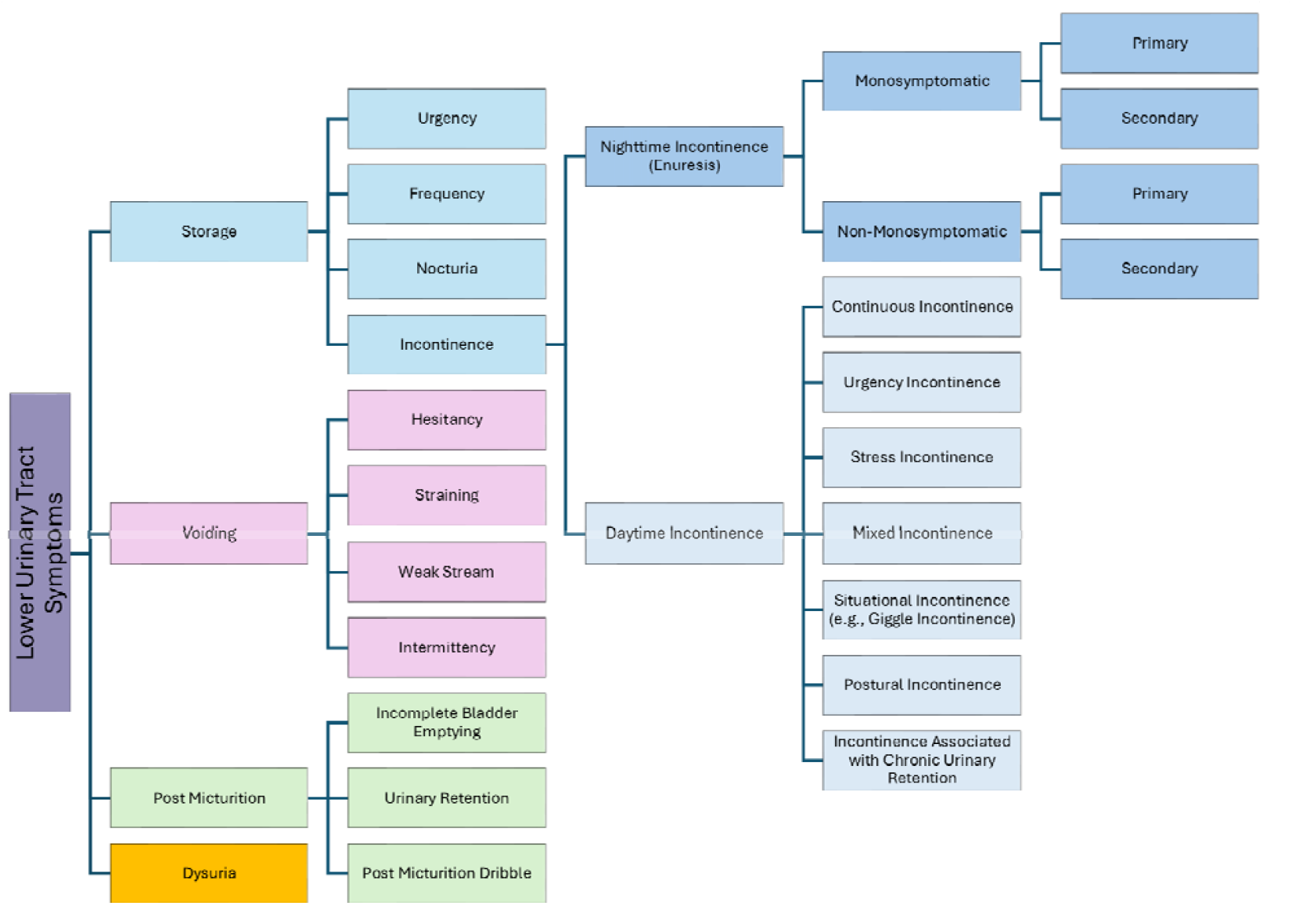
Subcategories of lower urinary tract symptoms

As individuals age from childhood to adolescence, LUTS may evolve, likely reflecting the physiological and hormonal developments characteristic of these growing years. In childhood, LUTS are primarily characterized by incontinence issues; however, neurologically intact children and adolescents may also exhibit voiding issues. These issues are commonly attributed to dysfunctional voiding, a condition in which the urethral sphincter or pelvic floor fails to relax during urination, leading to hesitancy, straining, prolonged micturition, and partial or complete interruptions in urine flow ^5^. As individuals advance into older adulthood, the diversity and prevalence of LUTS become more pronounced, and the range of issues significantly expands due to conditions such as benign prostatic obstruction (BPO) and stress urinary incontinence (SUI) in women, particularly during the postpartum period.

The implications of LUTS extend beyond physical symptoms, affecting psychological well-being and quality of life across the life course ^6–15^. LUTS are often reported alongside anxiety symptoms, suggesting a potential psychosomatic relationship that merits further investigation. Among children, physical and social challenges, such as social isolation and teasing, can lead to lowered self-esteem and distress. Additionally, the persistence of LUTS may exacerbate stress and can severely impact daily functioning and mental health.

The prevalence of LUTS is estimated to be 20-30% among adults and changes with aging ^16,17^, with significant variances due to different definitions and diagnostic criteria. Among children, approximately 7-10% consult specialists for LUTS, such as urinary frequency or dysuria due to recurrent infections and/or urinary incontinence ^18^. However, broader LUTS are reported by up to 25% of children, with common manifestations including daytime incontinence, holding maneuvers, and urinary urgency ^19^. The habitual delay of urination in children, using holding maneuvers, is classified as voiding postponement which often lead to low frequency, urgency and potentially incontinence. Voiding postponement is associated with psychological comorbidity in children ^20,21^.

Overactive bladder syndrome (OAB) is characterized by excessive urgency and frequency, affecting about 5-12% of children aged 5-10 years, many of whom continue to experience symptoms into adulthood ^22^. Population-based studies indicate OAB prevalence is 10.8% in men and 12.8% in women ^23–25^. These studies reveal sex-specific differences in symptoms, underscoring the importance of tailored management strategies to address the unique needs of each sex and various age groups ^26^.

Despite the recognized prevalence of LUTS and their impact on quality of life, their population-based incidence, particularly among pediatric and young adult populations, remains largely unquantified. To address this significant gap in epidemiological data, we aimed to estimate the sex-specific incidence of LUTS in children and young adults using a population-based sample from Sweden. We have developed an algorithm based on information sourced from the Swedish national registers, enabling a more accurate and comprehensive identification of LUTS cases.

## 2. Material and methods

### 2.1 Study population

We used data from the Swedish national registers to estimate the sex-specific incidence (sex assigned at birth) of LUTS. The comprehensive Swedish healthcare system, which encompasses all residents, facilitates the systematic collection of healthcare data through national registers.

Each individual is assigned a unique personal identity number, enabling the linkage of their health information across various registers ^27^.

Our study population consisted of liveborn singleton children born between January 1, 1995, and December 31, 2005, who were recorded in the Swedish National Medical Birth Register ^28^, with complete maternal and paternal information. We had access to follow-up data for these individuals until December 2018 or migration from Sweden. The National Medical Birth Register, operational since 1973, captures birth and neonatal data for approximately 98% of all births in Sweden. The reliability and completeness of the National Medical Birth Register data are assured through semi-automated extractions from electronic health records and rigorous quality checks by the National Board of Health and Welfare (Socialstyrelsen) ^28^.

Ethical approval and waiver of informed consent (which is not required for the use of register data) were obtained from the Regional Ethical Review Board in Stockholm, Sweden.

### 2.2 Outcomes and Exposure Covariate

We obtained diagnostic information about LUTS from the Swedish National Patient Register. The National Patient Register includes the Swedish National Inpatient Register (also known as slutenvårdsregistret or the Hospital Discharge Register), established in 1964 and achieving nationwide coverage by 1987. The National Inpatient Register accurately registers over 99% of all somatic and psychiatric hospital discharges in Sweden. Since 2001, Swedish counties have been obligated to report hospital-based outpatient physician visits to the National Patient Register. Although the coverage of inpatient care in the National Inpatient Register is almost complete, the coverage for outpatient care data is less comprehensive, mainly due to the partial inclusion of data from private healthcare providers ^29^. Additionally, since 2006, there has been a consistent decrease in the proportion of missing primary outpatient diagnoses ^30^. Nonetheless, it is important to recognize that variations in the sensitivity and completeness of the National Patient Register data still exist. Quality control is actively conducted by the National Board of Health and Welfare, which includes efforts to correct suspected errors by requesting new data from practitioners ^30^. This quality assurance process is essential in mitigating some of the limitations in data coverage and enhancing the overall accuracy of the register.

Diagnoses within the National Patient Register are made by clinical specialists, at inpatient and outpatient care, and are recorded using the International Classification of Diseases (ICD) codes. This practice ensures the accuracy and consistency of diagnostic information. However, it is crucial to note that diagnoses made in primary care settings are generally not included in the National Patient Register. This exclusion is significant for our study, as it implies that LUTS cases managed or diagnosed in primary care may not be directly represented in analyses. The absence of primary care data underscores the need to interpret our findings within the context of the available data’s scope and coverage. This limitation also motivates us to extract LUTS cases using criteria beyond ICD diagnostic codes.

To address this gap, we supplemented the National Patient Register data with information from the Swedish Prescribed Drug Register, which records all medications dispensed throughout Sweden. This approach allows us to potentially identify individuals who may have received a diagnosis of LUTS in primary care, even when such diagnoses are absent from the National Patient Register, based on medication type. By integrating these data sources, we aimed to provide a more comprehensive view of LUTS prevalence and management across different healthcare settings.

We established three distinct criteria to define LUTS for our study, ensuring a broad and accurate capture of potential cases: (1) ICD-10 diagnostic codes: LUTS were defined using specific ICD-10 codes. For voiding symptoms, we included ICD code R33.9, and for storage symptoms, we used N39.4A, N39.4C, and R35.9B. We excluded individuals with neurogenic bladder from this LUTS category, as their conditions do not align with the definitions of LUTS used in our research. Consequently, the LUTS status for these individuals was adjusted to reflect this exclusion. (2) Broad ICD-Medication: This approach cross-referenced the Swedish Prescribed Drug Register, defining LUTS as either an ICD diagnosis or at least one instance of prescribed medication specific to LUTS management. This included alfa 1 receptor inhibitors for managing voiding symptoms and anticholinergics and Beta-3 receptor agonists for storage symptoms. This broader criterion helps capture cases where medication is prescribed, perhaps based on symptoms treated in primary care settings, where diagnoses might not be formally recorded in the National Patient Register. (3) Narrow ICD-Medication: this definition more stringent requires an ICD diagnosis of LUTS or at least two prescriptions of the aforementioned medications. These prescriptions must be dispensed more than 76 days apart but within one year. The 76-day interval was chosen to accommodate the common practice of patients refilling their prescriptions approximately 14 days prior to completing a typical 90-day cycle. This criterion is intended to identify more persistent or confirmed cases of LUTS where consistent medication use indicates ongoing management.

### 2.3 Statistical analysis

To estimate the sex-specific incidence rates of LUTS, we calculated the number of new LUTS cases per 100,000 person-years for male and females. We specifically stratified these incidence rates by sex to reflect the distinct patterns of LUTS. To test the differences in incidences between sexes, we applied the exact test of the ratio between two Poisson rates. The computation of person-years began from the date of birth and continued until the earliest of the following events: first LUTS diagnosis, emigration, death, or the end of follow-up in December 2018. Confidence intervals (CIs) for the total population and sex-specific incidence rates were calculated under the assumption of a Poisson distribution. The cumulative incidences of LUTS were estimated as (1 – the Kaplan-Meier estimator). Differences between estimated cumulative incidence curves were tested using the log-rank test.

Additionally, to determine the age at which LUTS incidence was highest, we calculated age-and sex-specific incidence rates. This calculation involved summing the number of LUTS cases within each age group across all birth year cohorts and determining the total number of person-years for each age group. The incidence rate for each age group was then derived by dividing the total number of cases by the total person-years.

All statistical analyses were performed using Statistical Software R.

## 3. Results

### Cohort Demographics and Baseline LUTS Incidence

Our study analyzed a cohort consisting of 1,013,744 individuals born in Sweden between 1995 and 2005. After applying exclusions, which included 30,840 non-singleton births and one case with missing sex data, the final cohort size was narrowed down to 982,903. Using only ICD-10 diagnostic codes, we identified 633 individuals (0.1%) diagnosed with storage LUTS and 1,116 individuals (0.1%) with voiding LUTS (Table 1). The incidence rate for storage LUTS was 3.5 per 100,000 person-years [95% CI: 3.2 - 3.8], and for voiding LUTS, 6.2 per 100,000 person-years [95% CI: 5.8 - 6.5].

**Table 1.** Prevalence and Incidence Rates of LUTS Among Individuals Born in Sweden from 1995-2005 (N=982,903)

|  | Total population |  |  | Male |  |  | Female |  |  | P-value |
| --- | --- | --- | --- | --- | --- | --- | --- | --- | --- | --- |
|  | Without LUTS<br>n (%) | With LUTS<br>n (%) | Incidence rate<br>#cases/100000<br>person-years | Without LUTS<br>n (%) | With LUTS<br>n (%) | Incidence rate<br>#cases/100000<br>person-years | Without LUTS<br>n (%) | With LUTS<br>n (%) | Incidence rate<br>#cases/100000<br>person-years |  |
| Storage LUTS<br>ICD-10 | 982,270 (99.9) | 633 (0.1) | 3.5 [3.2 - 3.8] | 505,189 (99.9) | 274 (0.1) | 2.9 [2.6 - 3.3] | 477,081 (99.9) | 359 (0.1) | 4.1 [3.7 - 4.5] | <0.001 * |
| Storage LUTS<br>Narrow ICD-Medication | 974,078 (99.1) | 8,825 (0.9) | 49.0 [48.0 - 50.0] | 500,631 (99.0) | 4,832 (1.0) | 52.2 [50.8 - 53.7] | 473,447 (99.2) | 3,993 (0.8) | 45.6 [44.2 - 47.0] | <0.001 * |
| Storage LUTS<br>Broad ICD-medication | 978,009 (99.5) | 4,894 (0.5) | 27.1 [26.4 - 27.9] | 502,633 (99.4) | 2,830 (0.6) | 30.5 [29.4 - 31.7] | 475,376 (99.6) | 2,064 (0.4) | 23.5 [22.5 - 24.6] | <0.001 * |
| Voiding LUTS<br>ICD-10 | 981,787 (99.9) | 1,116 (0.1) | 6.2 [5.8 - 6.5] | 504,816 (99.9) | 647 (0.1) | 7.0 [6.4 - 7.5] | 476,971 (99.9) | 469 (0.1) | 5.3 [4.9 - 5.8] | <0.001 * |
| Voiding LUTS<br>Narrow ICD-Medication | 980,905 (99.8) | 1,998 (0.2) | 11.1 [10.6 - 11.6] | 504,364 (99.8) | 1,099 (0.2) | 11.8 [11.1 - 12.6] | 476,541 (99.8) | 899 (0.2) | 10.2 [9.6 - 10.9] | 0.001 * |
| Voiding LUTS<br>Broad ICD-Medication | 980,905 (99.8) | 1,215 (0.1) | 6.7 [6.4 - 7.1] | 504,759 (99.9) | 704 (0.1) | 7.6 [7.0 - 8.2] | 476,929 (99.9) | 511 (0.1) | 5.8 [5.3 - 6.3] | <0.001 * |

\*A p-value of less than 0.05 was considered statistically significant.

### Extended Definitions of LUTS

Using the broad ICD-medication definition significantly increased the identification of individuals with LUTS. Specifically, this criterion led to the identification of 8,825 individuals (0.9% of the cohort) with storage LUTS and 1,998 individuals (0.2% of the cohort) with voiding LUTS. The corresponding incidence rates were notably higher than those determined by the ICD-10 definitions alone: 49.0 per 100,000 person-years [95% CI: 48.0–50.0] for storage LUTS and 11.1 per 100,000 person-years [95% CI: 10.6–11.6] for voiding LUTS (Table 1).

In contrast, the narrow ICD-medication definition, which likely captures more chronic cases, identified somewhat fewer individuals: 4,894 cases (0.5% of the cohort) with storage LUTS and 1,215 cases (0.1% of the cohort) with voiding LUTS. The incidence rates for this more restrictive criterion were 27.1 per 100,000 person-years [95% CI: 26.4–27.9] for storage LUTS and 6.7 per 100,000 person-years [95% CI: 6.4–7.1] for voiding LUTS (Table 1).

### Sex Differences in LUTS

For storage LUTS, incidence rates varied by sex and diagnostic criteria (Table 1). Per ICD-10 diagnostic codes, females had a significantly higher incidence rate than males (P < 0.001). However, using broad and narrow ICD-medication definitions, males showed slightly higher rates than females.

In contrast, voiding LUTS consistently showed higher incidence rates in males across all definitions. Using ICD-10 diagnostic codes, the rate for males was 7.0 per 100,000 person-years, compared to 5.3 for females (P < 0.001). This finding was consistent using both broad and narrow ICD-medication definitions.

### Developmental Trends in the Incidence of LUTS Across Sex and Age

Across all age groups, the broad ICD-medication definition consistently yielded higher cumulative incidences of both storage and voiding LUTS compared to the narrow and ICD-10 definitions (Figure 2). This pattern was observed in both sexes, with males generally exhibiting slightly higher cumulative incidences than females. In all cases, the cumulative incidence curves differed significantly between sexes (P < 0.001), except for the broad voiding LUTS definition (P = 0.001).

**Figure 2.**
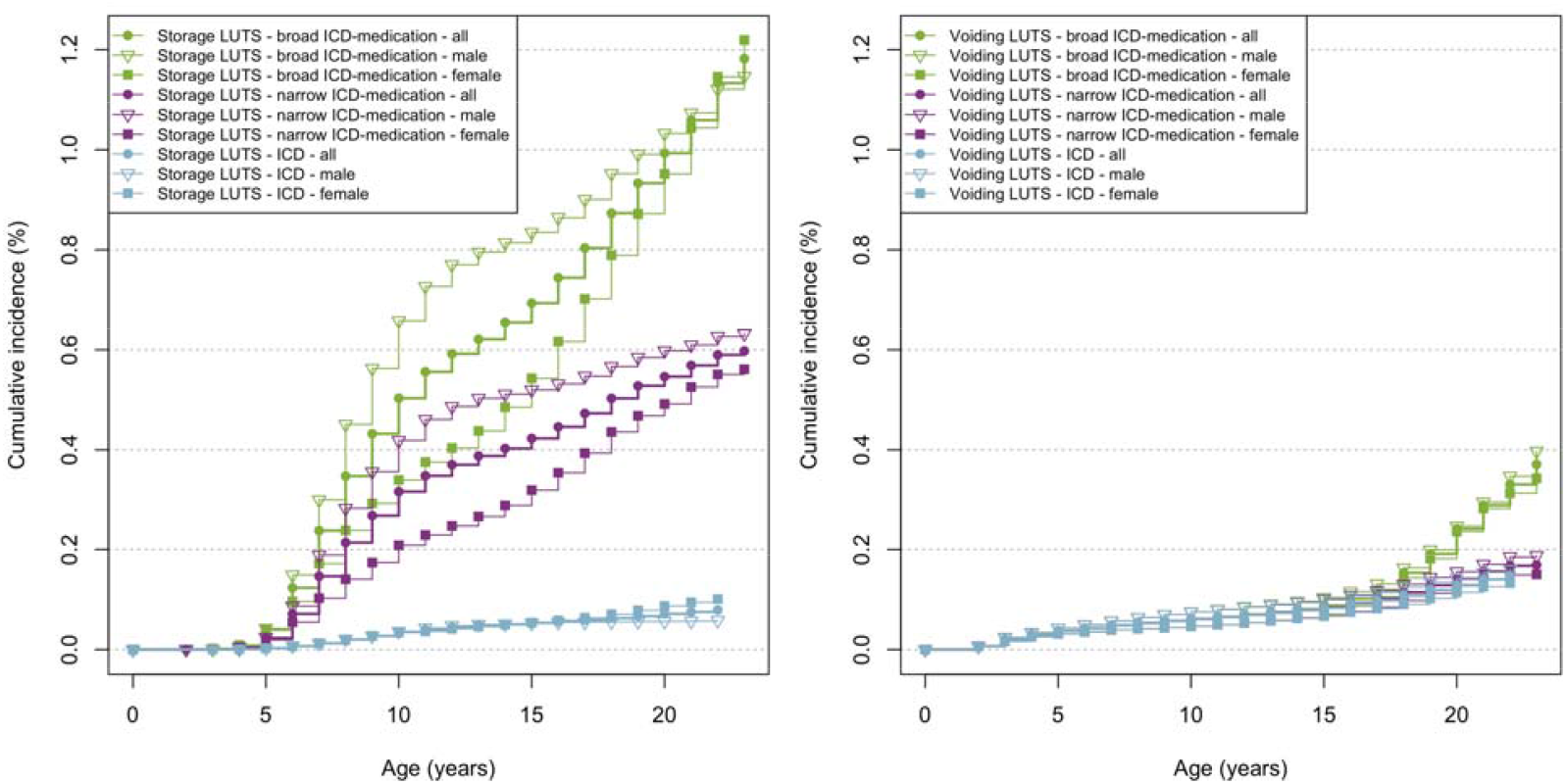
Cumulative incidence of storage LUTS (left) and voiding LUTS (right) under all three diagnostic criteria, for the entire population and stratified by sex. Green lines represent the broad ICD-medication definition, purple lines represent the narrow ICD-medication definition, and blue lines represent the ICD-10 definition. Solid circles denote all individuals, with markers differentiating between male (upward triangles) and female (squares) subjects.

### Diagnosis Age Differences Between Males and Females for LUTS

Significant sex differences were observed in the age at which LUTS was diagnosed, with males consistently diagnosed at younger ages than females across all definitions of both storage and voiding LUTS (Table 3). This pattern was statistically significant for each diagnostic criterion (P < 0.05), indicating a persistent sex-related difference in diagnosis timing.

**Table 3.**
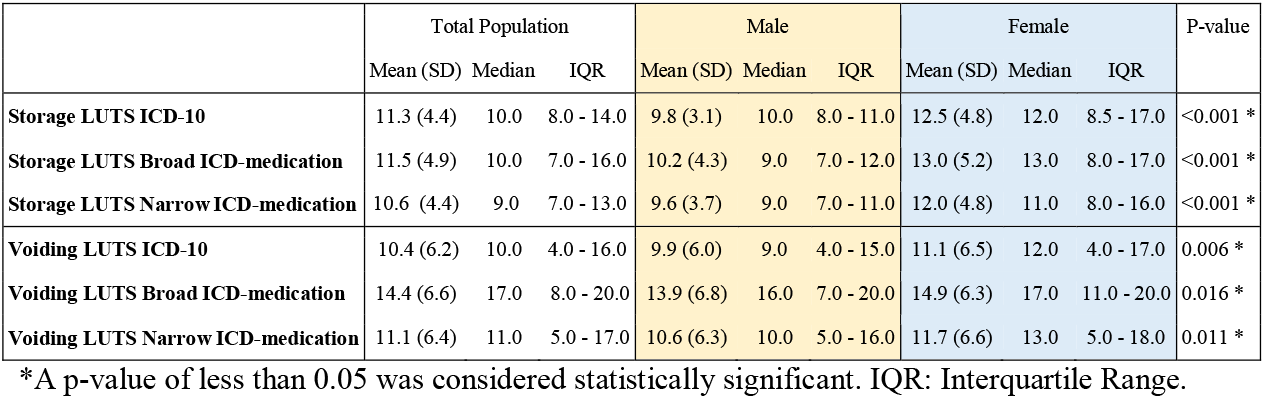
Diagnosis age of LUTS.

|  | Total Population |  |  | Male |  |  | Female |  |  | P-value |
| --- | --- | --- | --- | --- | --- | --- | --- | --- | --- | --- |
|  | Mean (SD) | Median | IQR | Mean (SD) | Median | IQR | Mean (SD) | Median | IQR |  |
| Storage LUTS ICD-10 | 11.3 (4.4) | 10.0 | 8.0 - 14.0 | 9.8 (3.1) | 10.0 | 8.0 - 11.0 | 12.5 (4.8) | 12.0 | 8.5 - 17.0 | <0.001 * |
| Storage LUTS Broad ICD-medication | 11.5 (4.9) | 10.0 | 7.0 - 16.0 | 10.2 (4.3) | 9.0 | 7.0 - 12.0 | 13.0 (5.2) | 13.0 | 8.0 - 17.0 | <0.001 * |
| Storage LUTS Narrow ICD-medication | 10.6 (4.4) | 9.0 | 7.0 - 13.0 | 9.6 (3.7) | 9.0 | 7.0 - 11.0 | 12.0 (4.8) | 11.0 | 8.0 - 16.0 | <0.001 * |
| Voiding LUTS ICD-10 | 10.4 (6.2) | 10.0 | 4.0 - 16.0 | 9.9 (6.0) | 9.0 | 4.0 - 15.0 | 11.1 (6.5) | 12.0 | 4.0 - 17.0 | 0.006 * |
| Voiding LUTS Broad ICD-medication | 14.4 (6.6) | 17.0 | 8.0 - 20.0 | 13.9 (6.8) | 16.0 | 7.0 - 20.0 | 14.9 (6.3) | 17.0 | 11.0 - 20.0 | 0.016 * |
| Voiding LUTS Narrow ICD-medication | 11.1 (6.4) | 11.0 | 5.0 - 17.0 | 10.6 (6.3) | 10.0 | 5.0 - 16.0 | 11.7 (6.6) | 13.0 | 5.0 - 18.0 | 0.011 * |

\*A p-value of less than 0.05 was considered statistically significant. IQR: Interquartile Range.

### Incidence Rate Differences Between Males and Females Across Age Groups

The incidence rates of LUTS classified by ICD-10 criteria across different ages reveal distinct patterns (Figure 3 and Figures S1 and S.2 in the supplement). Storage LUTS incidence exhibits a peak in late adolescence (7 to 9 years of age) for the overall cohort, with female incidence rates surpassing those of males in later teenage years (Figures 3b and 3c). Similarly, the incidence of voiding LUTS increases with age, culminating in the highest rates during late adolescence, although males demonstrate more significant fluctuations across the age spectrum (Figures 3d-3f). These trends underscore sex-specific differences in the development and diagnosis of LUTS, with females tending to be diagnosed at older ages, especially in the case of storage LUTS.

**Figure 3.**
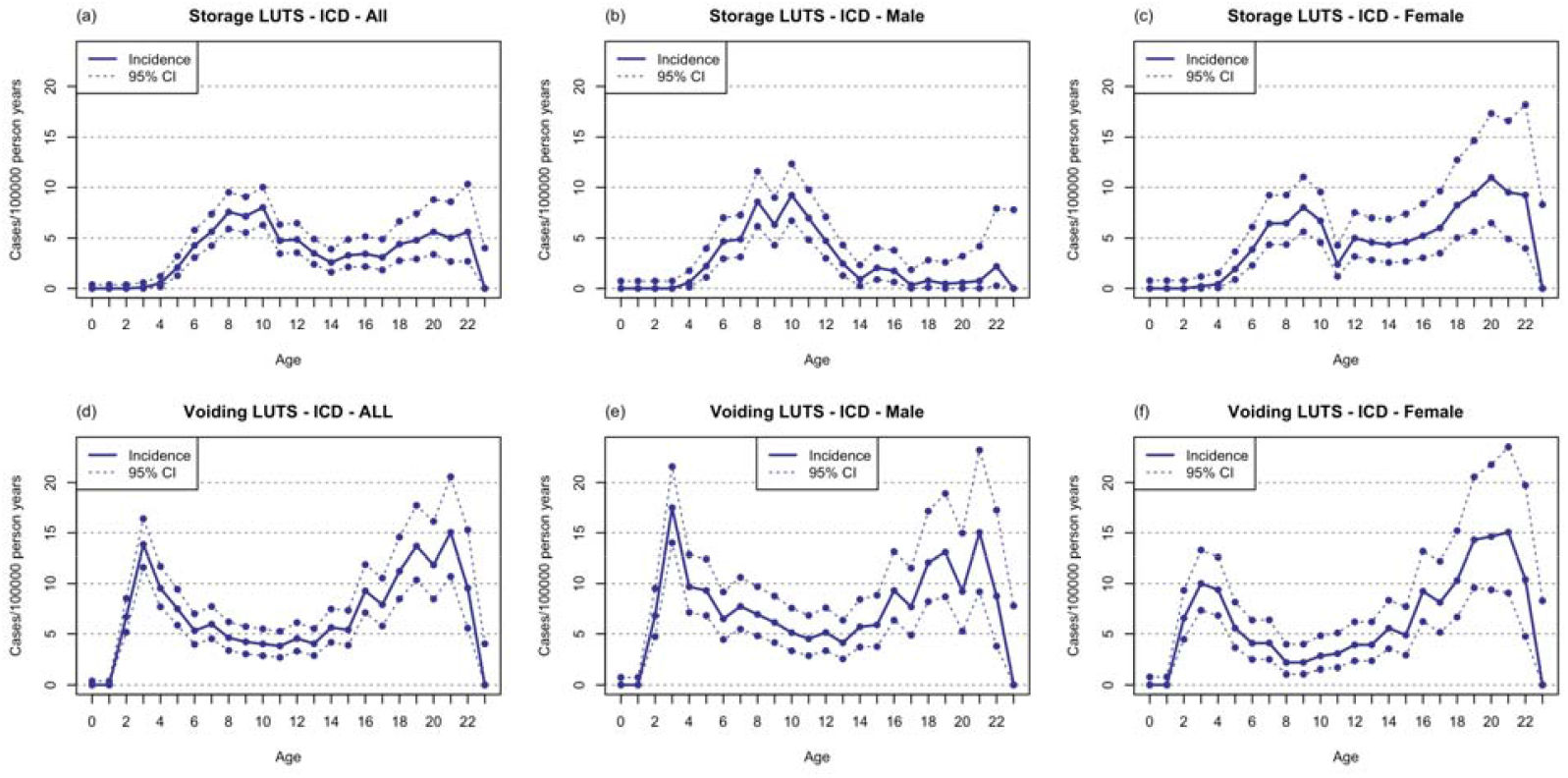
Developmental Trends in LUTS Incidence Across Sex and Age Groups. This figure illustrates the incidence rates per 100,000 person-years of storage LUTS (top row) and voiding LUTS (bottom row) across various ages, from infancy to early adulthood. Each column represents a different population group: the entire cohort (first column), male (second column), and female (third column). Solid lines indicate the estimated incidence, while dotted lines represent the 95% confidence intervals. These graphs highlight the developmental patterns of LUTS presentation and sex differences in incidence rates.

## 4. Discussion

This study analyzed a cohort of 982,903 Swedish individuals born between 1995 and 2005, and followed through 2018, using ICD-10 codes from the Swedish National Patient Register to identify LUTS. The initial findings revealed a low incidence rate of 0.1%—equivalent to 6.5 cases per 100,000 person-years for voiding LUTS and 3.7 cases per 100,000 person-years for storage LUTS—based solely on records from specialized hospital care records, which include inpatient and outpatient care but exclude primary and private care. This low rate suggests that LUTS diagnosed in hospital settings may represent a relatively small proportion of total cases.

When we expanded the diagnostic criteria to include both ICD codes and medication data from the Prescribed Drug Register, which covers prescriptions in all healthcare settings (including primary care), incidence rates increased substantially to 0.5% for storage LUTS (27.1 cases per 100,000 person-years) and 0.2% for voiding LUTS (11.1 cases per 100,000 person-years). This substantial increase highlights the limitations of relying solely on hospital-based ICD-10 coding and underscores the importance of integrating data from multiple healthcare sources to obtain a more accurate estimate of LUTS prevalence.

Further, our findings reveal notable sex differences in LUTS incidence rates, which varied depending on the diagnostic criteria applied. Males consistently showed higher incidence rates for voiding LUTS across all definitions. For storage LUTS, however, the pattern differed by criteria: using ICD-10 codes, females had a higher incidence rate (4.1 per 100,000 person-years) than males (2.9 per 100,000 person-years). In contrast, with broader and narrower ICD-medication definitions, males exhibited higher incidence rates for storage LUTS. Specifically, under the narrow ICD-medication criteria, the incidence was 30.5 per 100,000 person-years for males and 48.1 for females; under the broad criteria, the rates were 52.2 for males and 45.6 for females. These findings suggest that while females are more frequently diagnosed with storage LUTS under standard hospital-based coding, males have a higher incidence when medication data is included. This discrepancy may reflect differences in symptom management, potentially indicating that males are more frequently treated with medications for LUTS.

Moreover, our findings on age at diagnosis reveal a significant sex difference, with males typically diagnosed at younger ages than females. This difference may reflect an earlier onset of symptoms or greater symptom severity in males, or it could indicate more prompt recognition of symptoms by caregivers and healthcare providers in male patients.

This study has several strengths that set it apart from previous research on LUTS. First, our use of a large population-based sample minimizes selection bias, a common limitation in earlier LUTS studies that relied on small clinical samples of patients already diagnosed with LUTS, often alongside other pre-existing conditions. This approach reduces the likelihood of self-selection bias and provides a more representative cross-section of the general population. Additionally, the integration of multiple data sources, including the National Patient Register and the Prescribed Drug Register, significantly enhances the robustness of our findings. By capturing data from a broader range of healthcare settings, we can conduct a more accurate analysis of LUTS. This approach is especially valuable for individuals treated outside of hospitals, who might otherwise be missed in studies relying solely on specialty care data. The use of registry data spanning over a decade adds significant strength to our study, enabling us to examine trends in the incidence of LUTS across different age groups and over time. This approach provides valuable insights into the epidemiology of LUTS, including variations in diagnosis and management practices, offering a more comprehensive understanding of the condition than is possible with cross-sectional studies. Together, these strengths enable our study to provide a comprehensive and detailed assessment of population-based LUTS incidence rates, with particular focus on age and sex differences.

Register-based studies, while providing extensive data, have inherent limitations. A key issue is the potential time lag between the actual onset of a disorder and when patients seek medical care, which can lead to discrepancies in accurately capturing the timing and progression of LUTS. Additionally, reliance on diagnostic codes introduces variability, as coding accuracy depends on the healthcare provider’s familiarity with the system and clinical judgment. Misclassification bias may result if symptoms are incorrectly coded or if codes do not fully capture the patient’s condition, potentially leading to underestimation or overestimation of disease prevalence. Furthermore, many children and adolescents with LUTS receive conservative treatments, such as behavioral training, rather than medication. Consequently, our study may primarily capture patients with more pronounced LUTS symptoms.

In summary, this study’s use of a large population-based register has uncovered substantial underestimations in the incidence rates of LUTS, as well as significant sex and age disparities. Expanding the analysis to include broader diagnostic criteria and medication data underscores the need for improved diagnostic strategies and better integration of health records to accurately capture LUTS prevalence.

## Data Availability

Data from the Swedish national registers may be obtained from a third party and are not publicly available. Data cannot be shared publicly owing to restrictions by law. Data are available from the National Medical Registries in Sweden after approval by the Swedish Ethical Review Authority.

## Acknowledgments

This study was supported by a grant from the Swedish Research Council (2021-02846) and the Beatrice and Samuel A. Seaver Foundation, Icahn School of Medicine at Mount Sinai, New York, NY.

## Author Contributions

<u>Study concepts and design:</u> OA, JDB, DG, KH, BM, LRK, VS

<u>Acquisition, analysis, or interpretation of data</u>: OA, JDB, DG, KH, BM, LRK, VS

<u>Drafting of the manuscript:</u> All authors

<u>Critical revision of the manuscript for important intellectual content:</u> All authors

<u>Statistical analysis:</u> KH, BM, VS

<u>Obtained funding:</u> OA, BM

<u>Study supervision:</u> OA, BM

## Notes

### Competing Interest Statement

The authors have declared no competing interest.

