## Supplement for "Age- and Sex-Specific Prevalence of Lower Urinary Tract Symptoms in Pediatric and Young Adults in a Swedish National Cohort"

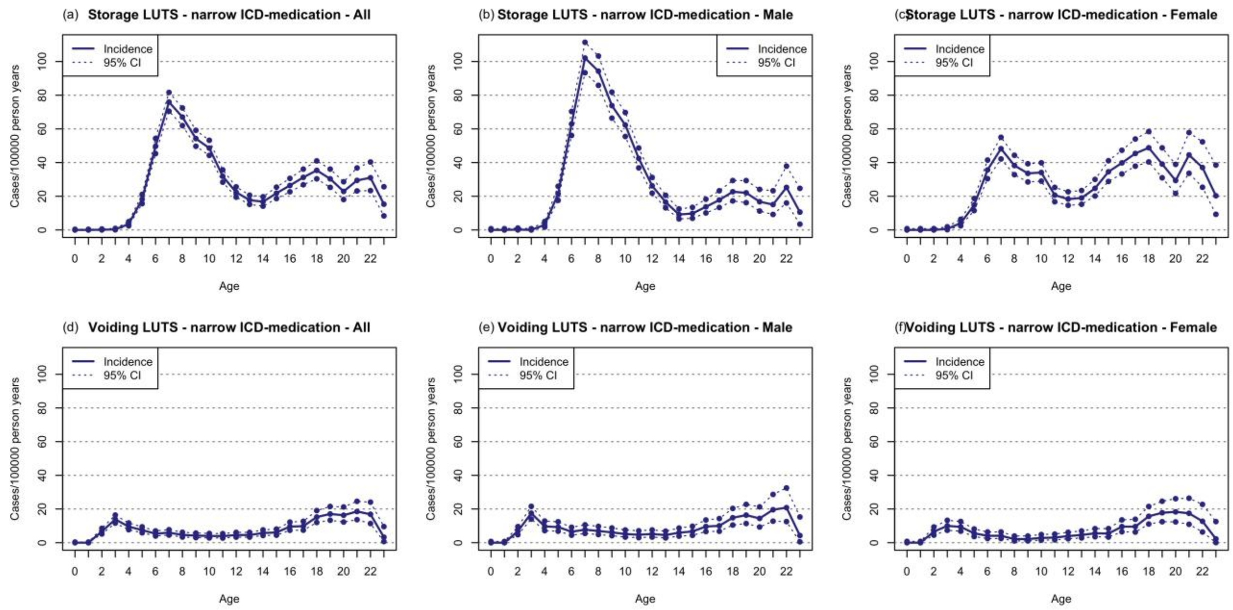


**Figure S1.** Developmental Trends in LUTS Incidence Across Sex and Age Groups, using narrow ICD-medication. This figure illustrates the incidence rates per 100,000 person-years of storage LUTS (top row) and voiding LUTS (bottom row) across various ages, from infancy to early adulthood. Each column represents a different population group: the entire cohort (first column), male (second column), and female (third column). Solid lines indicate the estimated incidence, while dotted lines represent the 95% confidence intervals. These graphs highlight the developmental patterns of LUTS presentation and sex differences in incidence rates.

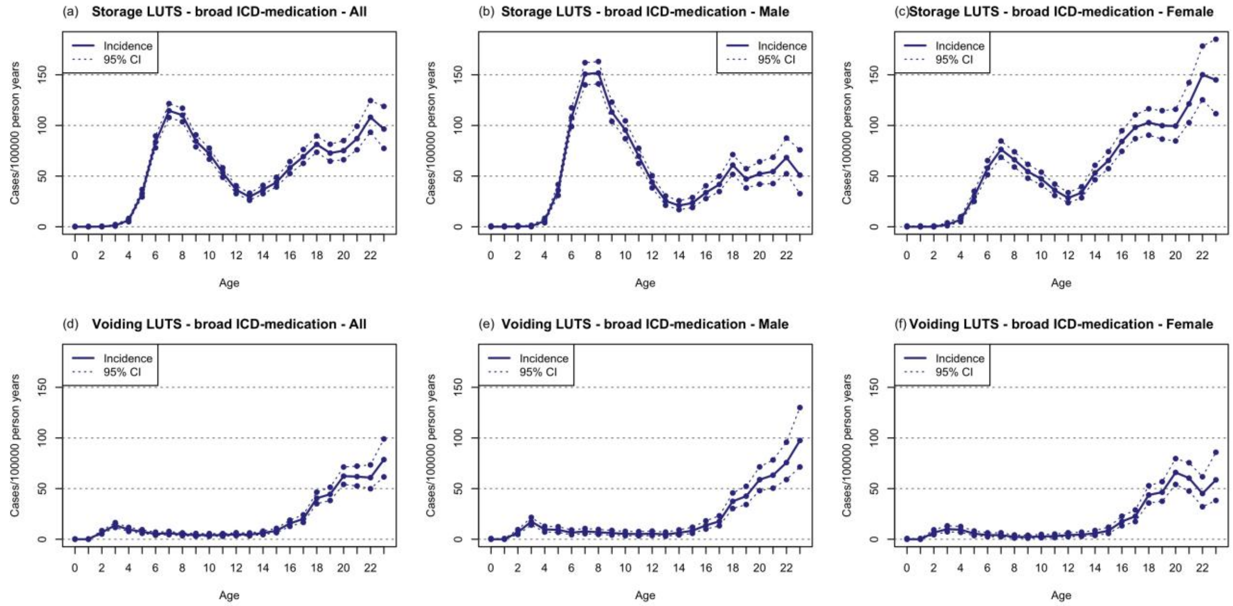


**Figure S2.** Developmental Trends in LUTS Incidence Across Sex and Age Groups, using broad ICD-medication. This figure illustrates the incidence rates per 100,000 person-years of storage LUTS (top row) and voiding LUTS (bottom row) across various ages, from infancy to early adulthood. Each column represents a different population group: the entire cohort (first column), male (second column), and female (third column). Solid lines indicate the estimated incidence, while dotted lines represent the 95% confidence intervals. These graphs highlight the developmental patterns of LUTS presentation and sex differences in incidence rates.
